# Patterns and Outcomes of Occupational Injuries in Emergency Departments at Nigerian Hydropower Plant Hospitals: A Retrospective Study

**DOI:** 10.64898/2026.08.31.26361543

**Authors:** John Olusola Ojo

## Abstract

Occupational injuries remain a cause of preventable morbidity and death, but evidence from emergency departments in Nigeria is limited. This retrospective study described occupational injuries treated at two hydropower plant hospitals between May 2020 and December 2022. Records of 120 patients were reviewed using a standardized abstraction form. Data were summarized using frequencies and percentages, and associations were assessed using chi-square tests. Vehicular crashes were the leading cause of injury (50.0%), lacerations were the most frequent injury type (39.2%), and the head/face was the most commonly affected single site (30.0%). Most patients were discharged (85.0%); 12.5% were transferred and 3.3% died. Serious injuries accounted for 40.8% of cases. Injury type was associated with discharge and injury severity, while injury site was associated with discharge and transfer. These findings support stronger workplace prevention, timely emergency assessment, and clearer referral pathways for injured workers.

## Introduction

Occupational injuries are an important cause of preventable illness, disability, and death, with consequences for workers, families, employers, and health systems.[1-3] Their burden is difficult to define in many low- and middle-income countries because occupational health coverage, surveillance, and reporting remain limited.[1,4,5] Emergency departments are important points of care for acute work-related injuries and provide information on injury mechanisms, anatomical sites, severity, and immediate outcomes.[6-9]

Published evidence linking occupational injury patterns with emergency care outcomes in Nigeria remains limited. This study described the causes, types, anatomical sites, severity, and immediate outcomes of occupational injuries presenting to two hydropower plant hospitals in Niger State, Nigeria, and examined associations between injury patterns and outcomes.

### Subjects and Methods

A retrospective descriptive study was conducted using emergency department records from two hydropower plant hospitals in Kainji and Jebba, Niger State, Nigeria. The hospitals served employees of the hydropower facilities and surrounding communities. Records were eligible if the patient had an injury classified as occupational and presented between May 2020 and December 2022. Records with incomplete information and injuries not related to work were excluded.

Of 523 emergency injury records screened, 161 were identified as possible occupational injuries. After excluding 41 incomplete or non-occupational records, 120 records were included. Data were extracted into a standardized Microsoft Excel spreadsheet. Variables included age, sex, marital status, occupation, health-care financing, cause of injury, injury type, anatomical site, discharge, transfer, death, length of hospital stay, and injury severity.

Injury severity was classified with the Abbreviated Injury Scale (AIS): minor (AIS 1), moderate (AIS 2), serious (AIS 3), severe (AIS 4), critical (AIS 5), and maximal or unsurvivable (AIS 6).[16] The study protocol and data collection tool were approved by the Ethics Committee of Cedarcrest Hospitals Limited, Niger State, Nigeria. Patient confidentiality and anonymity were maintained.

### Statistical Analysis

Data were analyzed using IBM SPSS Statistics for Windows, version 23.0 (IBM Corp., Armonk, NY, USA). Continuous variables were summarized using the mean and standard deviation, while categorical variables were reported as frequencies and percentages. Chi-square tests were used to assess associations between injury patterns and outcomes. A P value <0.05 was considered statistically significant.

## Results

The 120 patients had a mean age of 33.82 ± 13.02 years; 87 (72.5%) were aged 15-39 years, 104 (86.7%) were male, and 72 (60.0%) were married. Health insurance was recorded for 62 patients (51.7%).

Vehicular crashes were the most common cause of occupational injury (50.0%), followed by being hit by an object (32.5%) and falls, slips, or trips (15.0%). Lacerations were the most frequent injury type (39.2%), followed by multiple injuries (20.8%). The head/face was the most commonly affected single anatomical site (30.0%); 20.8% had injuries involving multiple body regions [Table 1].

**Table 1:** Patterns of occupational injury (N = 120)

| **Characteristic** | **n (%)** |
| --- | --- |
| Cause of injury |  |
| Vehicular crash | 60 (50.0) |
| Hit by object | 39 (32.5) |
| Fall/slip/trip | 18 (15.0) |

| Characteristic | n (%) |
| --- | --- |
| Heat/explosion | 2 (1.7) |
| Machine handling/lifting | 1 (0.8) |
| Type of injury |  |
| Laceration | 47 (39.2) |
| Multiple injuries | 25 (20.8) |
| Fracture | 13 (10.8) |
| Bruise/abrasion | 12 (10.0) |
| Traumatic brain injury | 11 (9.2) |
| Other* | 12 (10.0) |
| Anatomical site |  |
| Head/face | 36 (30.0) |
| Multiple regions | 25 (20.8) |
| Hand | 17 (14.2) |
| Lower limb/leg | 17 (14.2) |
| Other sites† | 25 (20.8) |

*\*Avulsion, chest injury, burn, dislocation, amputation, and crush injury. †Neck/spine, thorax/back, abdomen, upper arm/shoulder, forearm/wrist, and foot.*

Overall, 102 patients (85.0%) were discharged, 15 (12.5%) were transferred to another facility, and 4 (3.3%) died. Most patients, 116 (96.7%), had a recorded stay of 0-5 days. Serious injuries (AIS 3) were most frequent (40.8%), followed by moderate injuries (AIS 2; 39.2%) and severe injuries (AIS 4; 15.0%) [Table 2].

**Table 2:** Outcomes and injury severity (N = 120)

| **Outcome** | **n (%)** |
| --- | --- |
| Discharged | 102 (85.0) |
| Transferred | 15 (12.5) |
| Died | 4 (3.3) |
| Length of stay 0-5 days | 116 (96.7) |
| Length of stay 6-10 days | 2 (1.7) |
| Length of stay >10 days | 2 (1.7) |
| AIS 1: Minor | 2 (1.7) |
| AIS 2: Moderate | 47 (39.2) |
| AIS 3: Serious | 49 (40.8) |
| AIS 4: Severe | 18 (15.0) |
| AIS 6: Maximal/unsurvivable | 4 (3.3) |

*AIS: Abbreviated Injury Scale. Outcomes are presented as recorded in the source records and may not be mutually exclusive.*

Injury type was associated with discharge outcome (χ^2^ = 22.422, P = 0.013) and AIS category (χ^2^ z= 133.484, P < 0.001). Injury site was associated with discharge (χ^2^ = 21.878, P = 0.016) and transfer (χ^2^ = 21.475, P = 0.018). Cause of injury was not associated with discharge, transfer, death, or AIS category [Table 3].

**Table 3:** Associations between injury pattern and outcome.

| Comparison | $\chi^2$ | P value |
| --- | --- | --- |
| Cause of injury and discharge | 3.422 | 0.490 |
| Cause of injury and transfer | 3.604 | 0.462 |
| Cause of injury and death | 4.072 | 0.396 |
| Cause of injury and AIS category | 19.970 | 0.222 |
| Injury type and discharge | 22.422 | 0.013 |
| Injury type and transfer | 15.960 | 0.101 |
| Injury type and death | 12.113 | 0.278 |
| Injury type and AIS category | 133.484 | <0.001 |
| Injury site and discharge | 21.878 | 0.016 |
| Injury site and transfer | 21.475 | 0.018 |
| Injury site and death | 4.276 | 0.934 |
| Injury site and AIS category | 47.298 | 0.199 |

AIS: Abbreviated Injury Scale. $P < 0.05$ was considered statistically significant.

## Discussion

Vehicular crashes accounted for half of the occupational injuries in this study. This pattern differs from reports in which machinery, sharp objects, falls, or blunt trauma were the leading mechanisms.[6-8,10-12] Such variation is expected because occupational injury patterns reflect the industry, work tasks, environment, and route of presentation. The participating hospitals served hydropower facilities and surrounding communities, which may partly explain the prominence of transport-related events.

Lacerations were the most frequent injury type, while the head/face was the most common single anatomical site. Emergency department studies have also described lacerations and soft-tissue injuries as common presentations, although injuries to the upper or lower extremities predominate in several settings.[6-9,11,12] The differences reinforce the need for local surveillance rather than relying solely on findings from other industries or countries.

Most patients were discharged, although 12.5% required transfer and 3.3% died. Four in five injuries were classified as moderate or serious. High discharge proportions have also been reported in emergency department studies of occupational injury.[6,8,9,11,12] In this study, injury type was associated with discharge and AIS category, while injury site was associated with discharge and transfer. The clinical form and anatomical distribution of injury may therefore help identify patients who need a higher level of care, although the observed associations do not establish causality.

The study has limitations. It relied on retrospective records that were not created for research, and incomplete documentation may have affected classification. The findings came from two hospitals linked to hydropower communities and may not represent other Nigerian workplaces. Some categories contained few observations, which may reduce the reliability of chi-square testing. In addition, the recorded outcomes were not necessarily mutually exclusive and did not capture long-term disability or return to work. Despite these limitations, the study provides hospital-based evidence from a setting where published occupational injury data remain limited. Prevention should address transport safety, struck-by-object hazards, falls, machine safety, safety training, and appropriate use of personal protective equipment.[10,13-15] Emergency departments serving industrial communities should document work relatedness, injury mechanism, anatomical site, severity, disposition, and referral consistently. Larger prospective studies using standardized trauma measures are needed to examine predictors of death, transfer, disability, and return to work.

## Conclusion

Vehicular crashes, lacerations, and head/face injuries were prominent among occupational injuries treated at two Nigerian hydropower plant hospitals. Most patients were discharged, but transfers, serious injuries, and deaths were recorded. Injury type and anatomical site were associated with selected emergency care outcomes. Strengthening prevention, emergency assessment, documentation, and referral pathways may improve the care of injured workers.

## Data Availability

The individual-level analytical dataset used in the original study could not be retrieved for the present reanalysis. All aggregate cell counts used to reproduce the statistical analyses are reported in the Tables of the manuscript. No identifiable patient data are included.

## Financial Support and Sponsorship

Nil.

## Conflicts of Interest

There are no conflicts of interest.

## Data Availability Statement

The data supporting this study are not publicly available because they were derived from confidential hospital records. Requests may be considered by the corresponding author and the relevant institution, subject to ethical and institutional approval.

## Notes

### Competing Interest Statement

The authors have declared no competing interest.

### Author Declarations

The Ethics Committee (Kainji and Jebba Divisions) of Cedarcrest Hospitals Limited gave ethical approval for this work on 18 November 2022.

